# Subclinical SARS-CoV-2 Infections and Endemic Human Coronavirus Immunity Shape SARS-CoV-2 Saliva Antibody Responses

**DOI:** 10.1101/2024.05.22.24307751

**Authors:** Tonia L. Conner, Emilie Goguet, Hannah Haines-Hull, Allison Segard, Emily S. Darcey, Priscilla Kobi, Bolatito Balogun, Cara Olsen, Dominic Esposito, Milissa Jones, Timothy H. Burgess, Robert J. O’Connell, Christopher C. Broder, David Saunders, Simon Pollett, Eric D. Laing, Edward Mitre

## Abstract

This study characterized antibody responses induced by COVID-19 mRNA vaccination and SARS-CoV-2 infection in saliva. Utilizing multiplex microsphere-based immunoassays, we measured saliva anti-SARS-CoV-2 spike IgG, IgA, and secretory IgA in 1,224 saliva samples collected from healthcare workers in the Prospective Assessment of SARS-CoV-2 Seroconversion study between August of 2020 through December of 2022. By spring of 2022, most individuals had detectable spike-specific antibodies in saliva. Longitudinal measurements of saliva anti-SARS-CoV-2 nucleocapsid IgG revealed that most spike-specific IgA and secretory IgA detected in saliva was driven by subclinical and clinically-evident infections, rather than by vaccination alone. In contrast, saliva anti-SARS-CoV-2 spike IgG was strongly induced by vaccination and exhibited improved durability with hybrid immunity. Baseline levels of saliva antibodies to the endemic human coronaviruses positively correlated with post-vaccination anti-SARS-CoV-2 spike IgG levels. This study provides insights for development of vaccines that generate mucosal antibodies to respiratory pathogens.

**HIGHLIGHTS:** Saliva anti-spike antibodies were present in > 90% of participants by spring 2022

Saliva anti-spike IgA was driven by subclinical and clinically evident infections

COVID-19 mRNA vaccination alone was a weak inducer of saliva IgA antibodies

HCoV immunity correlates with post-vaccine anti-spike saliva antibody levels

## INTRODUCTION

Vaccines based on mRNA technology were the first approved for human use during the COVID-19 pandemic^1^ and their implementation led to substantial reductions in mortality and morbidity during the pandemic.^2^ However, while intramuscular (IM) mRNA vaccines reduced the severity of SARS-CoV-2 infection in vaccine naïve recipients, protection against infection is incomplete, lacks durability against infection, and is not transmission-blocking.^3–5^ There were also concerns that pre-existing immunity to the endemic human coronaviruses (HCoVs) could impair vaccine efficacy due to cross-reactivity with SARS-CoV-2.^6,7^

Effective vaccine platforms for SARS-CoV-2 interfere with critical elements of viral pathophysiology. SARS-CoV-2 initiates infection by binding angiotensin-converting enzyme 2 (ACE2) receptors expressed throughout the mucosal surfaces of the respiratory tract.^8^ The mucosal-associated lymphoid tissues (MALT), and specifically, the nasal-associated lymphoid tissues (NALT), produce the first adaptive immune response in the upper respiratory tract.^9^ After exposure to viral antigens, the NALT seeds tissues in the upper respiratory tract with mature B cells capable of rapidly producing antibodies into the nasal and saliva secretions.^9–12^ The predominant antibody generated at mucosal sites is immunoglobulin A (IgA), produced as monomers (most common in the serum) or dimers (most common in mucosal secretions).^13^ Dimeric IgA has been shown to neutralize the SARS-CoV-2 virus more potently than monomeric IgA^14^ and is moved across epithelial cells of the mucosa by the polymeric IgA receptor (pIgR), which is cleaved once reaching the apical side of the cell.^15^ Once the pIgR is cleaved, a 5-domain protein remains, referred to as the secretory component (SC). The SC stabilizes the dimer contributing to agglutination and exclusion of pathogens.^15^ When the SC is bound to dimeric IgA, the molecule is referred to as secretory IgA (SIgA),^16^ ^11,15^ and provides a total of four antigen-specific binding regions.^15^ The presence of dimeric IgA and SIgA in mucosal sites has been associated with early responses against viral pathogens, as well as lower viral load and shorter duration of viral shedding.^17,18^ Consequently, mucosal antibody responses are likely integral to protective immunity against SARS-CoV-2 infections.^19–22^

Understanding the predictors of mucosal immune responses in saliva and the kinetics of saliva antibody responses elicited by currently licensed IM COVID-19 mRNA vaccines is important for understanding vaccine control of viral shedding and transmission.^5,23^ This research is also central to the development of next generation COVID-19 vaccines which seek to block transmission and elicit robust mucosal immunity, including approaches which use a combination of IM and intranasal vaccines.^4^

While many studies have investigated circulating antibody levels in blood specimens after COVID-19 mRNA vaccination, relatively few have measured the effects mRNA vaccines have on saliva antibodies. Of those that have, most have found that COVID-19 mRNA vaccines induce spike-specific IgG antibodies in saliva.^24–29^ In contrast, results regarding induction of saliva IgA by mRNA vaccines have been diverse, with some reporting measurable increases in saliva IgA after initial or booster vaccinations^24–29^ and while other report minimal saliva IgA responses.^25,30,31^

One particular challenge in addressing the knowledge gap of how well IM mRNA vaccines induce saliva antibodies is accounting for subclinical infections that may confound vaccine study results. Thus, a key goal of this study was to determine how COVID-19 mRNA vaccination alone shapes mucosal antibody responses by identifying timepoints when study participants experienced clinical and subclinical SARS-CoV-2 infections and excluding these samples from longitudinal analyses. Another goal of this study was to determine whether pre-existing mucosal memory raised by prior endemic HCoV exposures influence saliva antibody responses induced by COVID-19 mRNA vaccination.

To achieve these goals, we characterized the patterns and predictors of longitudinal saliva antibody responses following COVID-19 mRNA vaccinations in a cohort of healthcare workers between August of 2020 through December of 2022. We incorporated evidence of both subclinical and clinical infections and observed remarkably different profiles of saliva antibody responses associated with infection alone, vaccination alone, and hybrid immunity. Additionally, we analyzed for predictors of vaccine elicited-saliva immunity by age, sex, and pre-vaccination (baseline) levels of saliva antibodies against the 4 major endemic HCoVs.

## METHODS

### Study Participants: Inclusion/Exclusion Criteria and COVID-19 testing

Participants were enrolled between August of 2020 and March of 2021 into the Prospective Assessment of SARS-CoV-2 Seroconversion (PASS) study, an ongoing prospective study evaluating clinical and immune responses to SARS-CoV-2 infection and vaccination. In-depth details of the PASS study methods have been published previously.^32^

At time of enrollment, all participants were employed at the Walter Reed National Military Medical Center (WRNMMC) and met the inclusion criteria of being ≥ 18 years of age and generally healthy. Participants were excluded at enrollment if they were immunocompromised, had history of COVID-19 prior to enrollment, or were SARS-CoV-2 seropositive. Additionally, for this analysis of saliva antibodies, participants were excluded from the analysis set if their initial COVID-19 2-dose vaccine series was not Pfizer BNT162b2.

All participants were asked to test for SARS-CoV-2, by either PCR testing at the WRNMMC COVID-19 testing facility and/or by rapid antigen testing at home, whenever they experienced symptoms consistent with possible SARS-CoV-2 infection. Participants reported vaccination and infection events at study clinic visits held monthly between August of 2020 and August of 2021, and then quarterly through December of 2022. Participants were classified as being uninfected until they either developed a positive SARS-CoV-2 test, or developed a doubling of their saliva IgG antibody levels against SARS-CoV-2 nucleocapsid (N) protein compared to their baseline level. Individuals that developed a doubling in saliva anti-SARS-CoV-2 (anti-SCV2) N IgG antibody levels, but had no history of a positive SARS-CoV-2 test, were categorized as having a “subclinical infection” based on a data-driven approach (see Results).

### Saliva Collection

Saliva samples were collected at quarterly clinic visits until August of 2022, and semi-annually since. While the PASS study is still ongoing, for this analysis we evaluated saliva samples collected through December 6, 2022. Whole saliva samples were collected by the passive drool method^33–36^ using the Salimetrics Saliva Collection Aid in accordance with the package insert (Salimetrics, State College, PA, USA). Participants were instructed to rinse their mouth with water for 10-15 seconds followed by a 10-minute rest period before collection. Saliva was then pooled in the mouth and drooled into the saliva collection device. Samples were frozen immediately after collection at -20°C, and then transferred later that day to -80°C for long term storage.

### Saliva antibody testing

On day of testing, saliva samples were thawed on ice and then centrifuged at 16,000 x g for 10 minutes at 4°C. Supernatant was transferred to a new tube, heat-inactivated for 30 minutes at 60°C, and then diluted 1:5 and 1:20 in phosphate buffered saline (PBS). Samples were tested for IgG, IgA, and SIgA using a well-described antigen-based multiplex microsphere immunoassay.^37–39^ Briefly, wild-type Wuhan-1 SARS-CoV-2 spike glycoprotein, expressed as native-like prefusion stabilized ectodomain trimer, hereafter referred to as ‘spike’, was sourced from Curia (Albany, NY, USA) and expression of trimeric ectodomain spike protein antigens of the four endemic human coronaviruses (HCoV-OC43, -HKU1, -NL63, -229E) have been previously described.^37,40^ The N protein was sourced from RayBiotech (Peachtree, GA, USA). S and N antigens (15 µg) were coupled to 100 µl of magnetic carboxylated beads (Luminex, Austin, TX, USA) and stored in accordance with the manufacturer’s protocol. A master mix of 1:100 beads to PBS ratio was prepared for each antibody plate. Antigen-coupled beads were incubated with 100 µl of each diluted saliva sample at room temperature for 45 minutes, under agitation at 700 rpm. Plates were then washed with PBS + 0.05% of Tween 20 a total of 3 times. Biotinylated cross-adsorbed goat anti-human IgG (Invitrogen, Waltham, MA, USA) was then added at a 1:5000 dilution for detection of IgG, and biotinylated cross-adsorbed goat anti-human IgA (Invitrogen) was used at a 1:5000 dilution for detection of IgA. Goat anti-human SIgA (MyBioSource, San Diego, CA, USA) was biotinylated using a biotinylation kit (Abcam, Cambridge, UK), and diluted at 1:5000 for SIgA detection. Plates were then incubated again at room temperature for 45 minutes, under agitation at 700 rpm. Plates were washed 3 times prior to incubation with a 1:1000 dilution of streptavidin-phycoerythrin (ThermoFisher, Waltham, MA, USA). After a final 3 washes, 100 µl of PBS + 0.05% of Tween 20 was added to each well and rocked at 700 rpm for 10 minutes prior to reading. Antigen-antibody levels were measured as a median fluorescence intensity (MFI) by a Bio-Plex 200 HTF multiplex system (BioRad, Hercules, CA, USA). Arbitrary binding units (AU/mL) for saliva antibodies were calculated by interpolating MFI values against in-house standard curves generated using microspheres conjugated with known concentrations of purified human IgG, IgA, or SIgA and serially diluted.

### Statistical Analysis

Comparisons were made using log-transformed data, with the Mann Whitney test for unpaired comparisons, and the Wilcoxon test for paired comparisons. The Kruskal-Wallis test followed by the Dunn’s test were used for multiple comparisons of three or more groups. Correlations were performed using Spearman’s rank correlation. Geometric means (GM), medians and interquartile ranges (IQR) were calculated using Graph Pad Prism Software version 10.

### Ethics

All participants provided informed consent and the protocol was approved by the Uniformed Services University Institutional Review Board in compliance with all applicable Federal regulations governing the protection of human participants.

## RESULTS

### Study demographics and vaccinations

The PASS study enrolled 271 participants from August of 2020 through March of 2021. Of these, 4 participants were excluded from this analysis: 1 participant was unable to provide saliva, 2 participants received the Moderna mRNA vaccine as their initial 2-dose vaccine series, 1 participant received the AstraZeneca vaccine, and 1 participant was found to be seropositive at baseline after repeat testing (Supplemental Figure 1). The self-reported demographics of the 266 included participants are presented in Supplemental Table 1. The majority were female (69.5%), white (71.4%), and non-Hispanic (92.1%), with a median age of 41 years (range 21-71; IQR 33.0-51.3). The most common occupations were nurse (32.7%), physician (25.9%), and occupational/physical/speech therapist (11.7%).

All participants who were vaccinated in this analysis received the Pfizer BNT162b2 WT spike COVID-19 mRNA vaccine as their initial 2-dose vaccination series (n=241). Of the 266 participants in the study, 22 only provided baseline samples and 2 participants did not get vaccinated. For the 3^rd^ vaccine dose, 163 participants received the BNT162b2 WT spike mRNA vaccine, and 2 received the Moderna WT spike mRNA-1273 vaccine. A total of 76 did not get a 3^rd^ dose during the study. Among the participants receiving a 4^th^ dose of the vaccine, 29 received the ancestral monovalent WT Pfizer BNT162b2 vaccine, 22 received the bivalent (WT+BA.4/5) Pfizer mRNA vaccine, and 1 received the bivalent (WT+BA.4/5) Moderna mRNA vaccine.

### Longitudinal anti-SCV2 spike and anti-SCV2 N saliva antibody levels

We evaluated the longitudinal kinetics of saliva IgG, IgA, and SIgA antibody responses to COVID-19 vaccination and SARS-CoV-2 infection over the course of the first 2 years of the pandemic (Figure 1). An increase in anti-SCV2 spike IgG was observed in the cohort around December of 2020 which aligned with the approval of the emergency use authorization for the first vaccines. By August of 2021, anti-SCV2 spike IgG levels in saliva declined, and continued to decline, until December of 2021 for those that had not yet received a 3^rd^ dose of the vaccine. Anti-SCV2 spike IgG levels increased in December of 2021 for the participants that received a 3^rd^ dose, but declined again by the following spring. Throughout 2022, we observed that most of the cohort had saliva anti-SCV2 spike IgG levels that remained comparable to levels at 1 month after the initial 2-dose vaccine series (Figure 1A).

**Figure 1.**
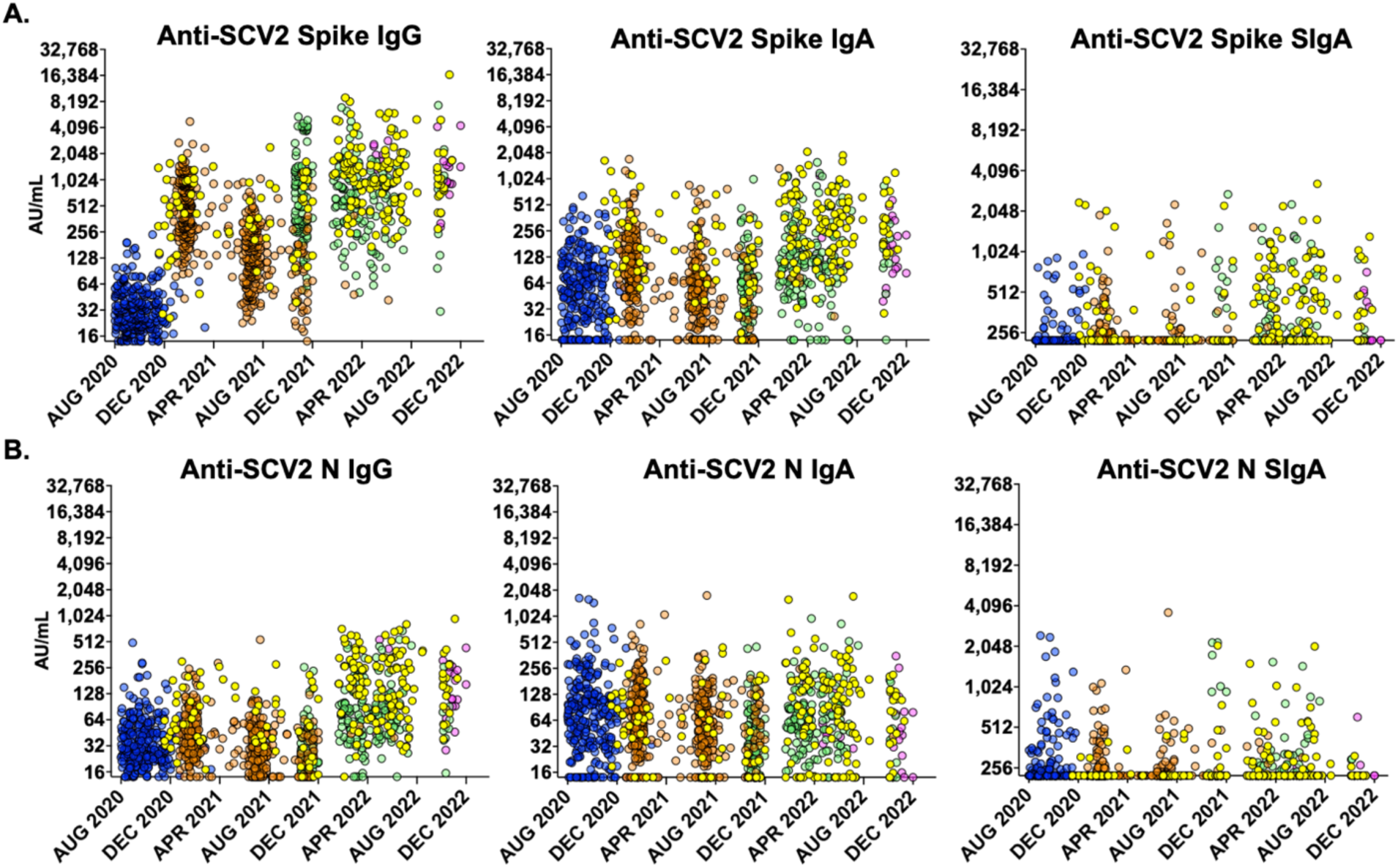
Longitudinal observations of antibody levels in saliva among PASS participants. **A)** Anti-SCV2 spike and **B)** anti-SCV2 N IgG, IgA and SIgA levels in saliva between August of 2020 through December of 2022 for all participants. Blue = baseline saliva samples (n=261, n=5 saliva quantity not sufficient for testing at baseline timepoint), orange = saliva samples after the 2^nd^ dose of mRNA vaccine (n=467), green = saliva samples after the 3rd dose of vaccine (n=263), pink = saliva samples after the 4^th^ dose of vaccine (n=30), yellow = saliva samples tested after documented SARS-CoV-2 infection (n=199)

In comparison, only modest increases in anti-SCV2 spike IgA levels were seen during the vaccine roll-out in late 2020 and throughout 2021. Substantially elevated levels of anti-SCV2 spike IgA were not consistently observed until 2022, by which time many in the cohort had confirmed infections and many others likely had subclinical infections. Similar dynamics were seen in anti-SCV2 spike SIgA (Figure 1A), with increases observed starting in April of 2022. Increases in anti-SCV2 N IgG were most consistently observed in 2022 (Figure 1B). Overall, few changes were observed in the levels of anti-SCV2 N IgA and SIgA in saliva over the course of the study (Figure 1B).

### Longitudinal assessment of subclinical SARS-CoV-2 infections

We suspected that the longitudinal increases in saliva anti-SCV2 spike IgA and SIgA levels observed in participants without a known positive test for SARS-CoV-2, were due to subclinical infections that increased in frequency during the pandemic.

To determine a saliva antibody threshold to categorize individuals with a possible subclinical infection, we evaluated anti-SCV2 N IgG levels in saliva from participants who developed post-vaccine SARS-CoV-2 infections (PVI) and compared their baseline levels of anti-N IgG to their levels 1-3 months after a PVI. Of these 68 participants with saliva samples collected between 1 and 3 months after a confirmed PVI, 59 participants exhibited saliva anti-SCV2 N IgG levels at least 2-fold greater than their baseline values, resulting in a sensitivity of 86.8% (Supplemental Figure 2).

On this basis, we then defined all participants who experienced at least a doubling in their saliva anti-SCV2 N IgG levels compared to baseline, as having a subclinical infection. Participants were thus ultimately categorized as follows: pre-vaccine infection (PreVI) (n=17), post-vaccine infection (PVI, n=73), subclinical post-vaccine infection (n=98), and no infection (n=59) (Supplemental Table 2). Because of participant attrition throughout the study, we next analyzed the percentage of participants that fit into each category at roughly quarterly intervals through December of 2022. By the fall of 2022, only 7.5% of participants evaluated had never tested positive for SARS-CoV-2 and never had saliva antibody evidence of a subclinical infection (Table 1).

**Table 1.** Vaccination and infection categorization of participants at specific time intervals.

| Date intervals | Number of participants tested at each interval | Number of participants that tested positive for SARS-CoV-2 at least once during the study | Number of participants with saliva anti-SCV2 N IgG doubling but no prior positive SARS-CoV-2 test (subclinical infection) | Number of participants with no prior positive SARS-CoV-2 test and no doubling of saliva anti-SCV2 N IgG (no prior infection) |
| --- | --- | --- | --- | --- |
| 8/25/2020 - 10/30/2020 | 181 | 0 (0.0%) | 0 (0.0%) | 181 (100.0%) |
| 11/1/2020 - 1/31/2021 | 112 | 10 (8.9%) | 13 (11.6%) | 89 (79.5%) |
| 2/1/2021 - 4/40/2021 | 192 | 13 (6.8%) | 65 (33.9%) | 114 (59.4%) |
| 5/1/2021 - 7/31/2021 | 158 | 8 (5.1%) | 60 (38.0%) | 90 (57.0%) |
| 8/1/2021 - 10/31/2021 | 64 | 9 (14.1%) | 22 (34.4%) | 33 (51.6%) |
| 11/1/2021 - 1/31/2022 | 163 | 16 (9.8%) | 65 (39.9%) | 82 (50.3%) |
| 2/1/2022 - 4/30/2022 | 138 | 48 (34.8%) | 59 (42.8%) | 31 (22.5%) |
| 5/1/2022 - 7/31/2022 | 135 | 61 (45.2%) | 57 (42.2%) | 17 (12.6%) |
| 8/1/2022 - 12/6/2022 | 53 | 24 (45.3%) | 25 (47.2%) | 4 (7.5%) |

### COVID-19 mRNA vaccinations induce detectable saliva levels of anti-SCV2 spike IgG antibodies but minimal anti-SCV2 spike IgA and SIgA antibody levels

Saliva antibody levels at specific timepoints after COVID-19 mRNA vaccination are shown in Figure 2. In analysis of all participants without a prior positive SARS-CoV-2 test at 1 month after the 2-dose initial vaccination series (1M PV2), we observed a substantial increase in anti-SCV2 spike IgG (15.2-fold increase compared to baseline), a modest increase in anti-SCV2 spike IgA (1.8-fold increase compared to baseline), and no measurable increase in anti-SCV2 spike SIgA (Figure 2A). Furthermore, we observed higher levels of anti-SCV2 spike IgG levels after the 3^rd^ vaccine dose (PV3) compared to the 2^nd^ vaccine dose (PV2). Additionally, the rate of decline in saliva anti-SCV2 spike IgG levels appeared to decrease after the 3^rd^ dose (PV3) compared to the 2^nd^ dose (PV2). We also observed no appreciable increase in anti-SCV2 spike IgA or SIgA at 1 month after the 3^rd^ dose (1M PV3) (Figure 2A). Because we observed increases in anti-SCV2 spike IgA and SIgA levels at later timepoints after the 3^rd^ vaccine dose (4-6M PV3 and 6-9M PV3), we posited that some participants were likely experiencing subclinical infections. In line with this, we also observed anti-SCV2 N IgG levels increase 2.4-fold in the 4-6M PV3 and 2.4-fold in the 6-9M PV3 groups (Supplemental Figure 3A).

**Figure 2.**
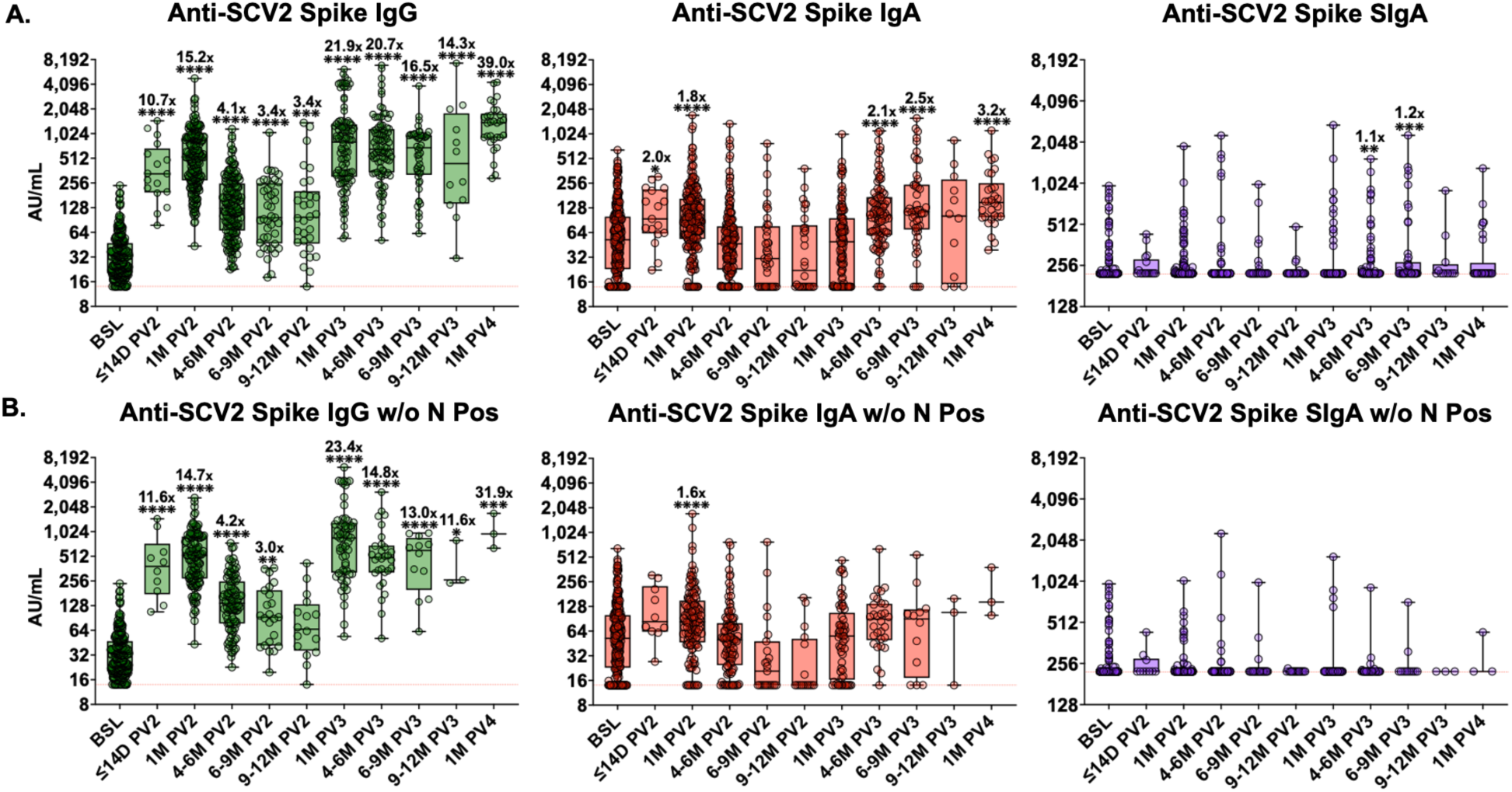
COVID-19 mRNA vaccinations induce detectable saliva levels of anti-SCV2 spike IgG antibodies but minimal anti-SCV2 spike IgA and SIgA antibody levels. **A)** Anti-SCV2 spike IgG, IgA and SIgA saliva antibody responses at each timepoint after vaccination compared to baseline levels in samples of participants with no prior documented SARS-CoV-2 infection (BSL n=261, ≤14D PV2 n=17, 1M PV2 n=205, 4-6M PV2 n=174, 6-9M PV2 n=41, 9-12M PV2 n=30, 1M PV3 n=110, 4-6M PV3 n=93, 6-9M PV3 n=49, 9-12M PV3 n=12, 1M PV4 n=30). **B)** Anti-SCV2 N IgG, IgA and SIgA saliva antibody responses at each timepoint after vaccination compared to baseline levels after removal of participants with a subclinical infection as well as those with prior documented SARS-COV-2 infection (BSL n=261, ≤14D PV2 n=10, 1M PV2 n=129, 4-6M PV2 n=104, 6-9M PV2 n=23, 9-12M PV2 n=16, 1M PV3 n=61, 4-6M PV3 n=30, 6-9M PV3 n=12, 9-12M PV3 n=3, 1M PV4 n=3). Comparisons made using log-transformed data and Kruskal-Wallis analysis with Dunn’s multiple comparison test. Fold-change in geometric mean compared to baseline levels is indicated in the text above box and whisker plots. * p < 0.05; ** p < 0.01; *** p < 0.001; **** p < 0.0001. Red dotted line represents lower limit of the assay. (BSL = baseline, D = days, M = months, PV2 = post-2^nd^ vaccine dose, PV3 = post-3^rd^ vaccine dose, PV4 = post-4^th^ vaccine dose).

Figure 2B shows saliva anti-SCV2 spike antibody responses to vaccination after removal of participants with known prior SARS-CoV-2 infection and participants with subclinical infections defined by doubling in anti-SCV2 N IgG. Overall, we observed only minor changes to anti-SCV2 spike IgA levels in saliva. A modest increase (1.6-fold increase compared to baseline) was observed in anti-SCV2 spike IgA levels in the 1M PV2 group, and the late (4-6M PV3 and 6-9M PV3) increases in anti-SCV2 spike IgA and SIgA were no longer present. In contrast, anti-SCV2 spike IgG levels remained markedly increased by vaccination with levels 14.7-fold greater than baseline at 1M PV2 and 23.4-fold greater at 1M PV3 (Figure 2B). These results demonstrate that while COVID-19 mRNA vaccination induces marked increases in saliva anti-SCV2 spike IgG, it induces only minimal increases in saliva anti-SCV2 spike IgA and SIgA.

### Boosting increases magnitude and durability of saliva anti-SCV2 spike IgG, but does not result in increases to saliva IgA or SIgA

To optimally compare the effect that additional vaccine doses have on saliva antibody levels, we removed saliva samples from participants with any prior positive SARS-CoV-2 test or evidence of prior subclinical infection. Anti-SCV2 spike IgG levels at 1 month after the 3^rd^ dose (1M PV3 = GM 747.1 AU/mL) were not statistically greater (p = 0.1579) than those obtained at 1 month after the 2^nd^ dose (1M PV2 = 469.1 AU/mL, Supplemental Figure 4). However, after a 3^rd^ dose, anti-SCV2 spike IgG levels in the 4-6M (4-6M PV3, GM = 471.4 AU/mL,) and 6-9M (6-9M PV3, GM = 413.3 AU/mL) groups were significantly greater (p < 0.0001 and p=0.0008, respectively) than those obtained at the same timepoints after initial 2-dose series (4-6M PV2 GM= 135.5 AU/mL and 6-9M PV2 GM = 95.31 AU/mL) (Figure 2B and Supplemental Figure 4). The trend showing improved durability over time in saliva anti-SCV2 spike IgG levels after a 3^rd^ dose was observed up to 9-12 months (9-12M PV3 vs 9-12M PV2, Supplemental Figure 4), though the comparison did not reach statistical significance. These differences were also observed using one-phase decay modeling. We determined the half-life of saliva anti-SCV2 spike IgG after 2 doses to be 92.7 days, 95% CI (71.1-121.5) while the half-life after 3 doses was 159.5 days, 95% CI (75.6-621.4) (Supplemental Figure 5). No appreciable differences were observed in anti-SCV2 spike IgA and SIgA levels at any timepoints after the 3^rd^ (PV3) and 4^th^ (PV4) doses (Figure 2B, Supplemental Figure 4).

### Females produce higher levels of vaccine elicited anti-SCV2 spike IgG and IgA in saliva than males, whereas antibody levels do not differ across age groups

To analyze saliva antibody levels between sexes and age groups at 1 month after a 2^nd^ vaccine dose (1M PV2), we again removed saliva samples from participants with any prior positive SARS-CoV-2 test or evidence of subclinical infection. Both anti-SCV2 S IgG and anti-SCV2 spike IgA levels were significantly greater in females compared to males (anti-SCV2 spike IgG GM = 525.4 AU/mL in females, GM = 364.5 AU/mL in males, p = 0.0023; anti-SCV2 spike IgA GM = 97.1 AU/mL in females, GM = 58.7 AU/mL in males, p = 0.0118, Figure 3A). No differences in saliva antibody levels were observed between age groups (Figure 3B).

**Figure 3.**
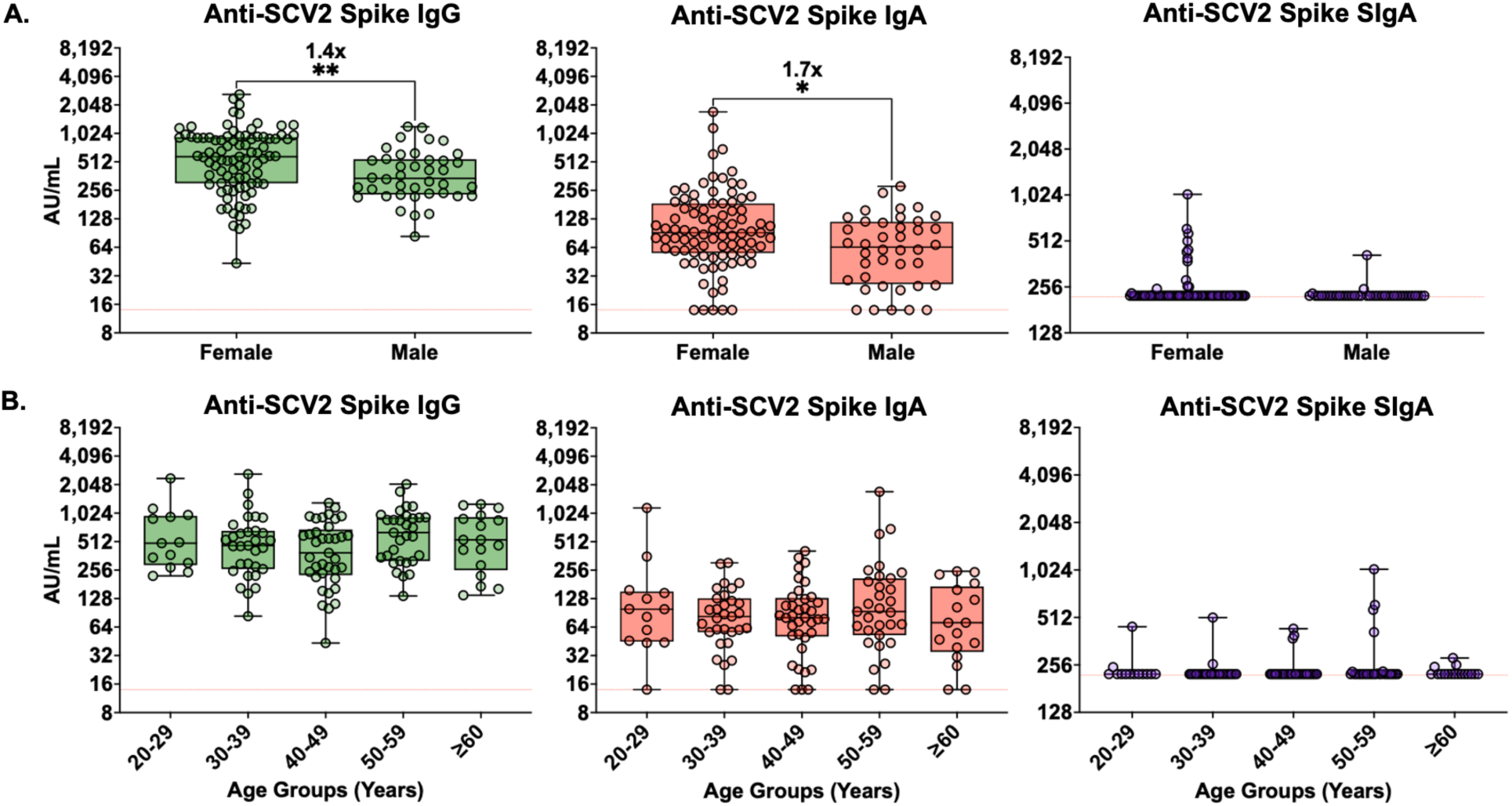
Females produce higher levels of vaccine elicited anti-SCV2 spike IgG and IgA in saliva than males, whereas antibody levels do not differ across age groups. A) Comparison of female (n=89) and male (n=40) saliva anti-SCV2 spike IgG, IgA and SIgA antibody responses 1 month after the 2^nd^ vaccine dose (1M PV2) in individuals with no prior documented or subclinical infection. Mann Whitney analysis performed on log-transformed data. Fold-change in geometric mean is indicated in the text above box and whisker plots. Red dotted line represents lower limit of the assay. B) Anti-SCV2 spike IgG, IgA and SIgA saliva antibody responses across age groups 1 month after the 2^nd^ vaccine dose (1M PV2) in individuals with no prior documented or subclinical SARS-CoV-2 infection. Comparisons made using log-transformed data and Kruskal-Wallis analysis with Dunn’s multiple comparison test. Red dotted line represents lower limit of the assay. (Age groups: 20-29, n=13; 30-39, n=31; 40-49, n=37; 50-59, n=31; 60 and above, n=17). * p < 0.05; ** p < 0.01; *** p < 0.001; **** p < 0.0001.

### Pre-vaccine infection induces significantly greater levels of anti-SCV2 spike IgA and SIgA in saliva than two doses of mRNA vaccine

To assess the effects SARS-CoV-2 infection had on saliva antibody levels in the absence of pre-existing immunity, we conducted paired analyses of saliva antibody levels in all participants that developed pre-vaccination infections (PreVI) and had saliva samples available both at baseline and 1 month after infection (n=14). Significant increases from baseline were observed in anti-SCV2 spike IgG levels with a 9.4-fold increase (p = 0.0002), anti-SCV2 spike IgA with a 4.4-fold increase (p = 0.0023), and anti-SCV2 spike SIgA with a 1.7-fold increase (p = 0.0039) (Figure 4A). A modest, but significant, increase (1.9-fold, p = 0.0203) was also observed in saliva anti-SCV2 N IgG levels while no increases were observed in saliva anti-SCV2 N IgA or SIgA in the pre-vaccine infection group (Figure 4B).

**Figure 4:**
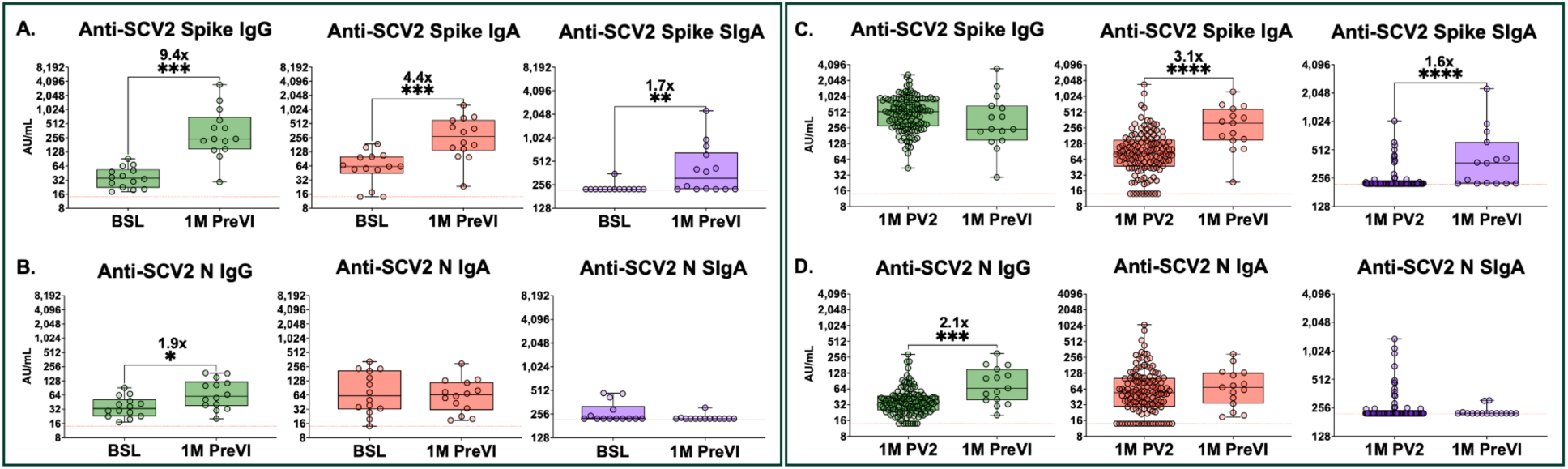
Pre-vaccine infection induces significantly greater levels of anti-SCV2 spike IgA and SIgA in saliva than two doses of mRNA vaccine. **A)** Anti-SCV2 spike and **B)** anti-SCV2 N IgG, IgA and SIgA responses in saliva at baseline (BSL) and 1 month after pre-vaccine infection (1M PreVI). Analysis performed on log-transformed data using Wilcoxson paired test (n=14). Fold-change in geometric mean compared to baseline levels is indicated in the text above box and whisker plots. * p < 0.05; ** p < 0.01; *** p < 0.001; **** p < 0.0001. Red dotted line represents lower limit of the assay. Comparisons of **C)** anti-SCV2 spike and **D)** anti-SCV2 N saliva antibody levels in saliva 1 month after 2^nd^ dose of vaccine (1M PV2, n=129) and 1 month after pre-vaccine infection (1M PreVI, n=15). All PV2 samples were from participants with no prior documented or subclinical SARS-CoV-2 infection. Analysis performed on log-transformed data using Mann-Whitney test. Fold-change in geometric mean is indicated in the text above box and whisker plots. * p < 0.05; ** p < 0.01; *** p < 0.001; **** p < 0.0001. Red dotted line represents lower limit of the assay.

We next compared saliva antibody levels measured 1 month after pre-vaccine infection (1M PreVI) with saliva antibody levels measured at 1 month after 2 vaccine doses (1M PV2) in individuals that had no prior positive SARS-CoV-2 test or evidence of subclinical infection. While vaccination and infection induced comparable levels of saliva anti-SCV2 spike IgG, the levels of anti-SCV2 spike IgA and SIgA were significantly greater after infection (anti-SCV2 spike IgA GM = 254.7 AU/mL in 1M PreVI, GM = 83.1 AU/mL in 1M PV2, p < 0.001; anti-SCV2 spike SIgA GM = 391.9 AU/mL in 1M PreVI, GM = 238.6 AU/mL in 1M PV2, p < 0.0001) (Figure 4C). As expected, saliva anti-SCV2 N IgG levels were greater after infection than after vaccination (Figure 4D).

### Hybrid immunity induces higher anti-SCV2 spike IgA and SIgA levels than 3^rd^ vaccine dose

To evaluate the impact hybrid immunity has on mucosal antibody levels, we compared saliva antibody responses in uninfected participants at 1 month after a 3^rd^ dose (1M PV3) to participants that had received 2 doses of vaccine and then developed a PVI (PV2+1M PVI) (Figure 5). A 3^rd^ dose induced the same saliva levels of anti-SCV2 spike IgG as 2 doses followed by a PVI. In contrast, anti-SCV2 spike IgA and SIgA levels were markedly higher in the hybrid immune group than in the 1M PV3 group. (anti-SCV2 spike IgA = 50.1 AU/mL in 1M PV3, and 328.3 AU/mL in PV2+1M PVI, p < 0.0001; anti-SCV2 spike SIgA = 246.0 AU/mL in 1M PV3, and 755.0 AU/mL in PV2+1M PVI, p < 0.0001) (Figure 5A). Anti-SCV2 N IgG and IgA levels were also significantly greater in the hybrid immunity group (Figure 5B).

**Figure 5:**
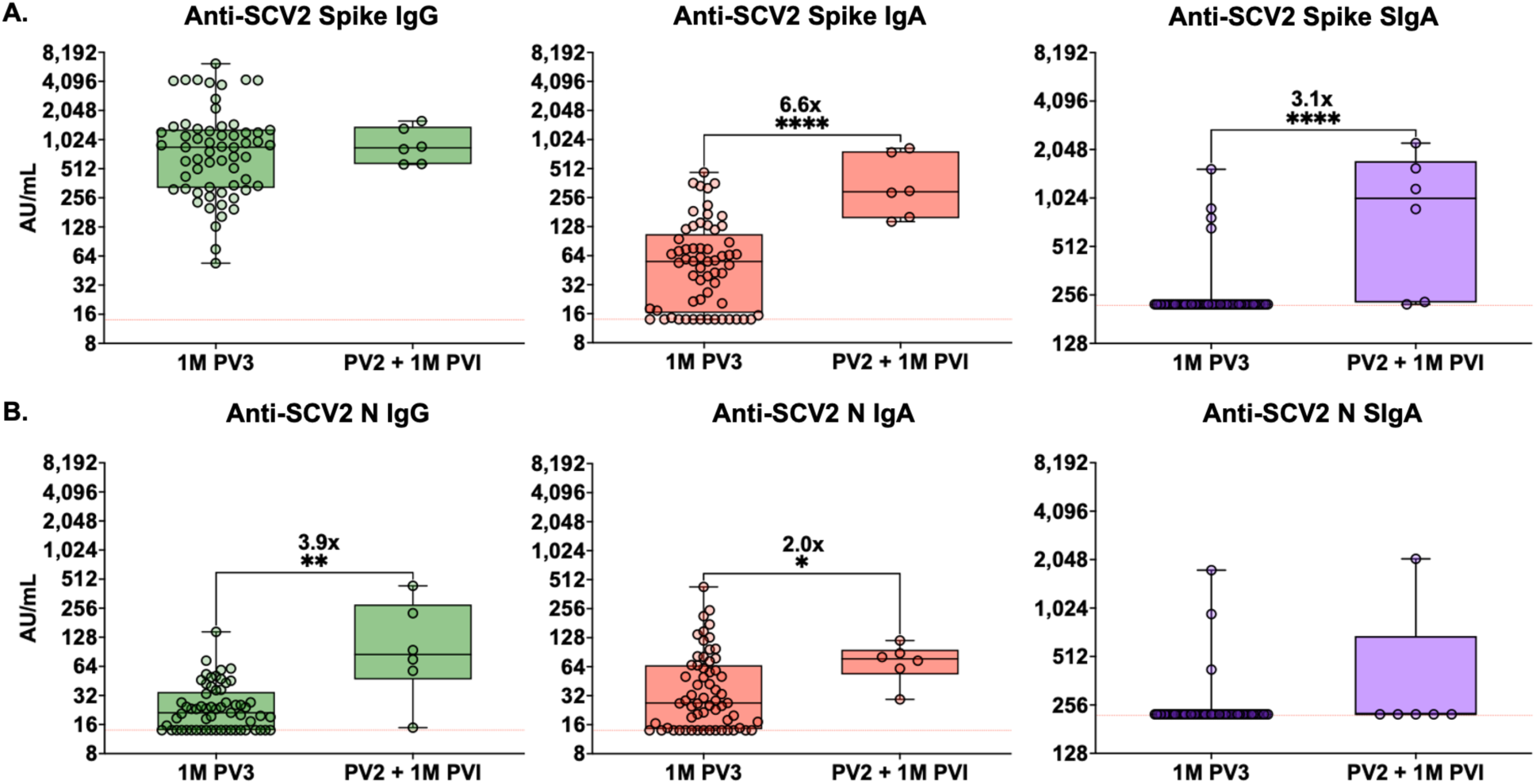
Hybrid immunity induces higher anti-SCV2 spike IgA and SIgA levels than 3^rd^ vaccine dose. **A)** Anti-SCV2 spike and **B)** anti-SCV2 N IgG, IgA and SIgA responses in saliva at 1 month after 3^rd^ vaccine dose in uninfected participants (1M PV2, n=61) compared with levels 1 month after infection in participants who received a 2^nd^ dose of vaccine and then had a post-vaccine infection (PV2 +1M PVI, n=6). Analysis performed on log-transformed data using a Mann-Whitney test. Fold-change in geometric mean is indicated in the text above box and whisker plots. ns, not significant; * p < 0.05; ** p < 0.01; *** p < 0.001; **** p < 0.0001. Red dotted line represents lower limit of the assay.

Additionally, we also examined hybrid immunity by comparing uninfected participants that received a 4th dose of vaccine (1M PV4) with participants that received a 3rd dose of vaccine and then had a PVI (PV3+1M PVI) (Supplemental Figure 6). Individuals with hybrid immunity (PV3+1M PVI) exhibited greater saliva IgA responses compared to individuals that had received a 4^th^ vaccine dose, but the differences were not statistically significant, likely due to the very small number of uninfected participants who received a 4^th^ dose of vaccine (1M PV4). Of note, both anti-SCV2 spike IgG and IgA levels in saliva were 3.6-fold and 2.2-fold greater, respectively, at 1 month after last antigen exposure in hybrid immune participants when vaccination was followed by infection (VVVI = 3 vaccine doses then infection) than when infection was followed by vaccination (IVVV = infection followed by 3 vaccine doses) (Supplemental Figure 7).

### Vaccination results in boosting of saliva IgG against the spike protein of OC43

Prior studies have shown that COVID-19 mRNA vaccines boost serum levels of IgG antibodies against the spike protein of OC43, one of the four HCoVs.^41–43^ To determine if a similar cross-reactive boosting phenomenon occurs in the saliva, we measured IgG, IgA, and SIgA levels in saliva against the spike proteins of OC43, 229E, HKU1, and NL63 at baseline and 1 month after vaccination (1M PV2) in participants with no prior positive SARS-CoV-2 test or subclinical infection. Vaccination resulted in a modest (1.3-fold) increase in IgG saliva levels against OC43 spike protein (p= 0.0006, Supplemental Figure 8). No statistically significant increases were observed in saliva antibody levels against the spike proteins of 229E, HKU1, and NL63. In participants that were unvaccinated and infected, a similar increase (1.4-fold) was observed 1 month after infection (1M PreVI) in anti-OC43 spike IgG, though the increase was not statistically significant (p = 0.3575, Supplemental Figure 9).

### Baseline saliva IgG levels against the spike proteins of all four HCoVs positively correlate with anti-SCV2 spike IgG saliva levels after vaccination

The BNT162b2 vaccine contains mRNA that codes for the WT SARS-CoV-2 spike protein. To determine if baseline mucosal immunity against HCoVs had an effect on vaccine-induced saliva antibody levels, we assessed for correlations between baseline saliva antibody levels to the spike proteins of the four HCoVs and 1-month post-vaccination (1M PV2) responses to the spike protein of SARS-CoV-2 (Figure 6). Again, this analysis was performed using samples from participants with no prior positive SARS-CoV-2 test or subclinical infection. We found saliva anti-SCV2 spike IgG levels after vaccination positively correlated with baseline saliva IgG levels against the S proteins of all four HCoVs. A modest correlation was found with baseline anti-OC43 spike IgG levels (Spearman’s rho = 0.4608, p < 0.0001), and weak correlations were observed with baseline IgG levels against spike proteins of 229E, HKU1, and NL63 (rho = 0.2719, p = 0.0021; rho = 0.2814, p = 0.0014; rho = 0.2872, p = 0.0011, respectively).

**Figure 6:**
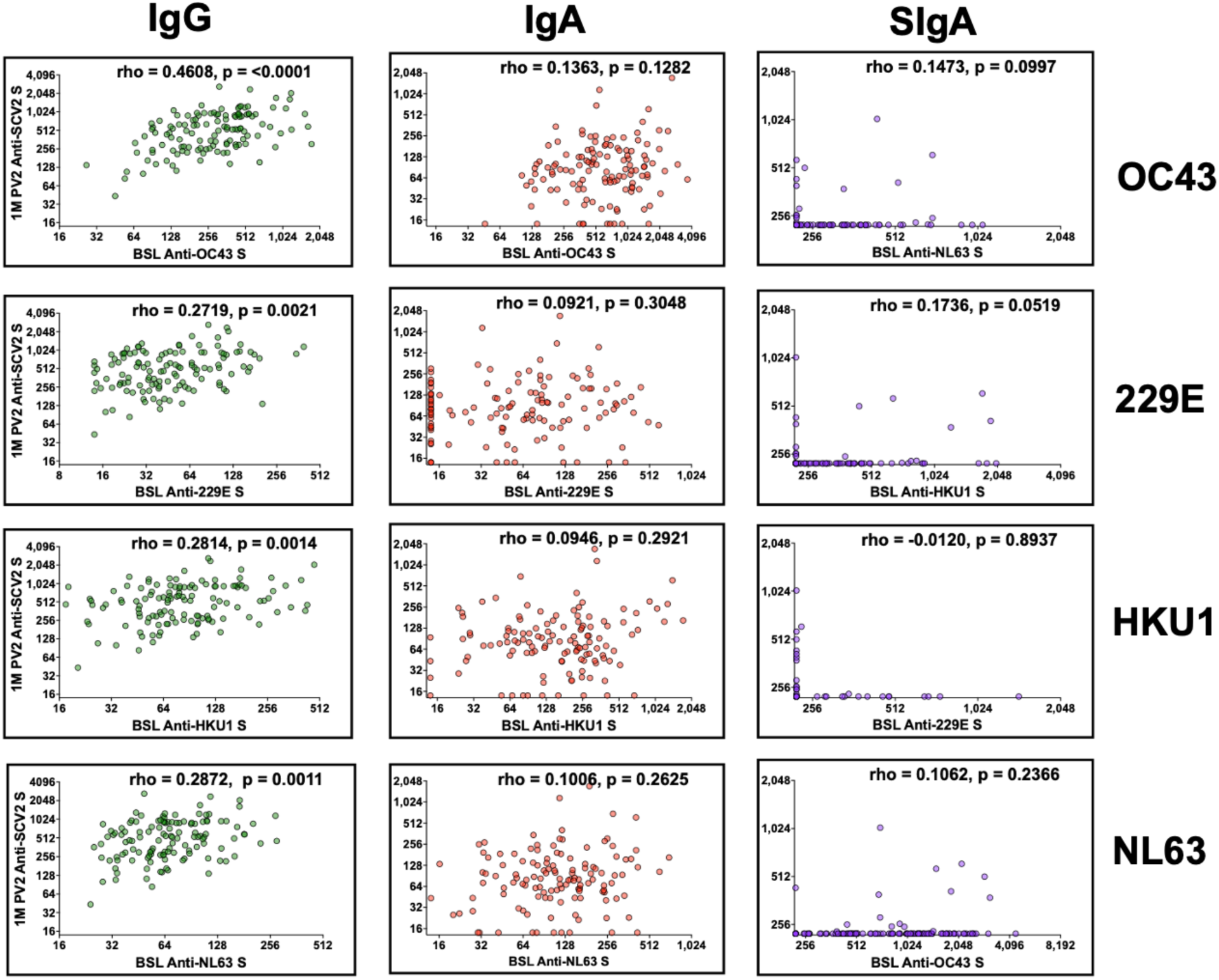
Baseline saliva IgG levels against the spike proteins of all four HCoVs positively correlate with anti-SCV2 spike IgG saliva levels after vaccination. Spearman correlations of paired baseline (BSL) saliva anti-spike (S) OC43, HKU1, 229E, and NL63 antibody levels with saliva anti-SCV2 spike (S) antibody levels 1 month after 2^nd^ COVID-19 mRNA vaccine (1M PV2, n=126). * p < 0.05; ** p < 0.01; *** p < 0.001; **** p < 0.0001. Red dotted line represents lower limit of the assay.

## DISCUSSION

In this prospective longitudinal analysis of 266 healthcare workers at the Walter Reed National Military Medical Center in Bethesda, Maryland, USA, we observed that by the spring of 2022 most individuals had detectable anti-SCV2 spike IgG and IgA antibodies in saliva. Our results indicate that the detection of anti-SCV2 spike IgA and SIgA in saliva was primarily driven by subclinical and clinically-evident infections. While COVID-19 mRNA vaccination alone was a weak inducer of saliva IgA antibodies, IgG was strongly induced in the saliva by vaccination and exhibited increased half-life after subsequent vaccinations and after a post-vaccine infection. Additionally, we observed that women generated greater levels of saliva IgG and IgA than men, and identified positive correlations between baseline saliva anti-HCoV spike IgG levels and anti-SCV2 spike IgG levels in saliva 1 month after vaccination.

A major challenge for evaluating how well COVID-19 mRNA vaccines induce mucosal antibodies is potential confounding by subclinical SARS-CoV-2 infections. By the summer of 2022, 45% of participants still being followed in the PASS study reported having at least 1 known SARS-CoV-2 infection. Because individuals can have pauci- or asymptomatic infection, especially in the post-vaccine era, we also sought to exclude all individuals with immunological evidence of a possible subclinical infection. This was done by eliminating, from vaccine-only analyses, all samples obtained after individuals exhibited a doubling of saliva anti-SCV2 N IgG antibodies compared to their baseline levels. Longitudinal analyses revealed that by the summer of 2022, only 12.6% of participants remaining in the cohort had no evidence of prior SARS-CoV-2 infection (45.2% with a known SARS-CoV-2 positive test and another 42.2% with saliva anti-SCV2 N IgG doubling without a prior positive SARS-CoV-2 test). This high rate of SARS-CoV-2 exposure is consistent with a Morbidity and Mortality Weekly Report stating that 70% of blood donors through September 2022 had either a pre-vaccine infection or post-vaccine infection as well as another study which estimated that 97% of the US population was infected by November 2022.^44,45^

Our study demonstrates that COVID-19 mRNA vaccination induces robust anti-SCV2 spike IgG levels in saliva but poor IgA responses in naïve individuals. After the initial 2-dose series, anti-SCV2 spike IgG antibodies were detectable in saliva as early as 14 days and peaked at 1 month to levels 14.7-fold greater than baseline. In contrast, anti-SCV2 spike IgA increased modestly, achieving a peak that was only 1.6-fold greater than baseline values, and anti-SCV2 spike SIgA never increased significantly above baseline.

We next evaluated how additional vaccine doses influenced the kinetics of saliva antibody levels. Our analysis of samples from infection naïve participants showed that a 3^rd^ vaccine dose resulted in 8.7-fold higher levels of saliva anti-SCV2 spike IgG than 2 doses. Additionally, a 3rd vaccine dose improved the durability of the saliva anti-SCV2 spike IgG response, with a half-life of 159.5 days determined by analyses performed through 9 months of follow-up. However, a 3^rd^ vaccine dose resulted in no appreciable increases in saliva anti-SCV2 spike IgA or SIgA.

While there are many publications examining antibodies in the serum after mRNA vaccination, studies of saliva have been less frequent. There is overall agreement in the literature that mRNA vaccination induces IgG responses in saliva.^24–29^ However, reports on saliva IgA responses after COVID-19 mRNA vaccines have varied. Several studies have reported detectable increases in saliva IgA after initial and/or booster vaccination with COVID-19 mRNA vaccines.^24,26–29,46^ Others, more consistent with our findings, have observed only minimal IgA responses in uninfected individuals.^25,30,31^

A potential limitation of COVID-19 vaccine studies is possible inclusion of individuals with prior subclinical infections, as vaccination after prior SARS-CoV-2 infection can induce significant saliva anti-SCV2 spike IgA responses.^47,48^ At least 2 studies that found increases in saliva IgA attempted to screen out unrecognized infections by testing for serum anti-SCV2 N IgG.^24,29^

While serum anti-SCV2 N is often positive shortly after infection, the sensitivity of anti-SCV2 N IgG for diagnosing prior infection is highly variable (52% - 100%)^49,50^ and its half-life post-infection is short, with one study estimating it to be only 35 days.^51^ In this study, we utilized saliva anti-SCV2 N IgG as a marker for possible prior infection. Saliva anti-SCV2 N IgG has been reported to have a high sensitivity for prior SARS-CoV-2 infections,^49^ and we found an 86% sensitivity for known post-vaccine infections in our cohort. Further, we obtained saliva antibody levels at baseline and then regularly (approximately 4 times a year), increasing the likelihood that we would not miss a transient infection-induced increase in saliva anti-SCV2 N antibodies. We believe the approach taken in our study convincingly demonstrates that COVID-19 mRNA vaccination (after the initial 2-dose series as well as after 3^rd^ or 4^th^ doses) induces only minimal increases in saliva IgA responses in the absence of prior infection. Of note, given that very few individuals remain that have not yet been infected at least once with SARS-CoV-2^44,45^, it will be challenging for future studies to investigate the impact COVID-19 vaccines can have on immunologically naïve individuals.

With regards to saliva antibody durability, a large cross-sectional study found that the anti-SCV2 RBD/spike IgG half-life in saliva was 140 days after 2 doses of mRNA vaccine.^52^ We found the half-life of saliva anti-SCV2 spike IgG to be 92.7 days after 2 doses of vaccine and 159.5 days after a 3^rd^ vaccine dose, analyzed through 9 months. Although reported half-lives in binding antibodies and neutralizing antibodies vary in the literature, the consensus is that a 3^rd^ dose does extend the durability of anti-SCV2 spike IgG in serum,^31,53–55^ which is also reflected in our saliva anti-SCV2 spike IgG data.

In addition to evaluating the effects of vaccine alone on saliva antibody responses, we also evaluated effects of pre-vaccine infection and post-vaccine infection. Pre-vaccine infection resulted in saliva IgG antibody levels that were similar to those induced by 2 doses of vaccine.

Additionally, post-vaccine infection (PV2 +1M PVI) resulted in saliva IgG antibody levels similar to those in naïve individuals after 3^rd^ vaccine dose (1M PV3). In contrast, anti-SCV2 spike IgA and SIgA were significantly greater after infection alone, and in the setting of hybrid immunity, compared to samples tested after vaccination alone, with the greatest values observed when infection occurred after vaccination. These results are consistent with other studies that have identified clear differences in humoral immune responses between vaccine-only groups and hybrid immune groups.^47,56,57^ These results suggest that development of anti-SCV2 spike IgA and SIgA in the saliva requires antigen exposure in the NALT to generate a local mucosal immune response through activation of tissue-resident memory B cells.

When COVID-19 vaccines were introduced, there was some concern that prior immunity to HCoVs could potentially impair vaccine-induced responses.^6,7^ To evaluate for this possibility, we conducted longitudinal measurements of saliva antibodies to the S proteins of the four HCoVs as well as to the spike protein of SARS-CoV-2. Rather than a negative effect, we observed a moderate positive correlation between baseline saliva anti-OC43 S IgG levels and saliva anti-SCV2 spike IgG levels measured 1 month after the initial 2-dose BNT162b2 vaccine series. We observed weak, but significant, positive correlations of post-vaccine saliva anti-SCV2 spike IgG with baseline saliva anti-SCV2 spike IgG levels to the HCoVs (229E, HKU1, and NL63). Anti-OC43 spike IgG levels in saliva were also boosted at 1 month after COVID-19 mRNA vaccination. The cross-reactivity of the spike proteins of OC43 and SCV2 has been shown in several studies of serum antibodies, and is consistent with their high sequence homology.^7,42,58–60^ Results of serological studies between other endemic HCoV IgG levels and vaccination response in the literature has been varied and many of these studies were conducted after infection rather than vaccination.^58,59,61–63^ Like our findings with saliva antibody levels, one serological study in humans found positive correlations between pre-existing antibody levels to the spike proteins of 229E and NL63 and post-vaccine levels of anti-SCV2 spike IgG.^59^ In contrast to observations in humans, two recent studies using mouse models observed that pre-existing immunity to HCoVs did not affect serum anti-SCV2 spike IgG responses to COVID-19 mRNA vaccination.^64,65^ An alternative explanation for the correlations we observed between HCoVs and SARS-CoV-2 vaccine response could be cross reactive anti-SCV2 spike IgG from a prior SARS-CoV-2 infection. We believe this to be unlikely however, since all participants included had negative IgG serology to the spike protein of SARS-CoV-2 upon enrollment in the study. Additionally, only participants without evidence of a subclinical infection, using the doubling of anti-SCV2 N IgG threshold, were used in the HCoV correlation analyses.

With regards to demographics, we observed that females produced higher saliva anti-SCV2 spike IgG and IgA than males after vaccination, a finding that has also been observed in studies of humoral immunity.^52,66^ In contrast, we found no effect of age on saliva antibody responses 1 month after vaccination which differs from serum studies which have found reduced antibody responses after COVID-19 mRNA vaccination in older populations.^67–69^ We speculate our study may not have observed differences with age because the maximum age was only 71 years old and participants with major comorbidities were excluded from the study.

### Limitations

Limitations of our study include small sample sizes for uninfected/unexposed participants at 9-12 months after the 3^rd^ dose and after the 4^th^ dose. Another limitation is that in this study we did not assess virus neutralization or other functional effects of saliva antibodies. However, in a recent publication by our group,^38^ we found that antibody from serum functioned as well as neutralizing antibodies as correlates of protection.^70^ Our study also was predominantly comprised of white females and was a single center study, both of which may impact the generalizability of our results.

### Conclusions

Our results support the current literature highlighting the need for further research on the mucosal immune system and its activation through vaccination. While IM mRNA vaccinations have reduced the burden of COVID-19, they have also highlighted gaps in our knowledge regarding approaches that can be taken to optimize mucosal immunity. Further research is warranted in this area to improve our ability to protect individuals against viral respiratory pathogens.^71–74^

## Data Availability

All data produced in the present study are available upon reasonable request to the authors.

## ACKNOWLEDGEMENTS

The authors gratefully acknowledge all PASS study participants.

The authors wish to also acknowledge all who have contributed to the PASS study:

Uniformed Services University (USU) of the Health Sciences: Christopher Broder, Timothy H. Burgess, Si’Ana Coggins, Tonia L. Conner, Emily S. Darcey, Anuradha Ganesan, Emilie Goguet, Hannah Haines-Hull, Belinda Jackson-Thompson, Eric D. Laing, Alyssa Lindrose, Edward Mitre, Matthew Moser, Robert J. O’Connell, Simon Pollett, Marana Rekedal, Mark Simons, David Tribble

Uniformed Services University Translational Medical Center: Heidi Adams, Bolatito Balogun, Milissa Jones, Priscilla Kobi, Lakeesha Kosh, Raquel Martinez, Roshila Mohammed, David Saunders, Dutchabong Shaw

Henry M. Jackson Foundation, Inc.: Kimberly Blankenship, Julian Davies, Mark Fritschlanski, Tigiste Girma, Luca Illinik, Orlando Ortega, Sanjeev Oudit, Edward Parmelee, Jennifer Rothenberg, Mimi Sanchez, Marianne Spevak, Todd Stroberg

Leidos Biomedical Research, Inc., Frederick National Laboratory for Cancer Research: John H. Powers, III

Naval Medical Research Center-Clinical Trials Center: Yolanda Alcorta, Christopher A. Duplessis, Monique Hollis-Perry, Andrew Letizia, Santina E. Maiolatesi, Elaine Morazzani, Kathleen F. Ramsey, Anatalio E. Reyes, Greg Wang, and Mimi A. Wong.

Walter Reed National Military Medical Center: Wesley Campbell, Sara Robinson

## AUTHOR CONTRIBUTIONS

Formal Analysis, T.L.C., C.O. E.M.; Writing-original draft, T.C.; Methodology, T.L.C., E.G., H.H, E.S. D., M.J., D.S., E.D.L., E.M.; Investigation, T.L.C., E.G., H.H., A.S., E.S.D, P.K., B.B.; Visualization, T.L.C.; Conceptualization, T.L.C., E.G., C.C.B, S.P., E.L., E.M.; Project Administration, E.G., M.J., D.S., T.H.B., R.J.O., S.P., E.D.L., E.M.; Writing-review & editing, T.L.C., E.G., M.J., D.S., T.H.B., C.C.B., R.J.O., S.P., E.D.L., E.M.; Resources, H.H., E.G., M.J., D.S., R.J.O., S.P., E.L., E.M.; Data curation, T.L.C., E.G., H.H., S.P., E.M.; Validation, T.L.C., E.S.D., E.D.L.; Supervision, E.G., M.J., D.S., R.J.O., S.P., E.D.L., E.M.; Funding acquisition, T.H.B., R.J.O., S.P., E.M.

## FUNDING

The protocol was executed by the Infectious Disease Clinical Research Program (IDCRP), a Department of Defense (DoD) program executed by the Uniformed Services University of the Health Sciences (USUHS) through a cooperative agreement by the Henry M. Jackson Foundation for the Advancement of Military Medicine, Inc. (HJF). This work was supported in whole, or in part, with federal funds from the Defense Health Program (HU00012020067, HU00012120094) and the Immunization Healthcare Branch (HU00012120104) of the Defense Health Agency, United States Department of Defense, and the National Institute of Allergy and Infectious Disease (HU00011920111), under Inter-Agency Agreement Y1-AI-5072, by the Armed Forces Health Surveillance Division (AFHSD), Global Emerging Infections Surveillance (GEIS) Branch, under award ProMIS ID P0099_22_NM and Navy WUN A1417, and by the US Food and Drug Administration Medical Countermeasures Initiative grant # OCET, 2022-1750. The sponsors had no involvement in the study design, the collection of data, the analysis of data, the interpretation of data, the writing of the report, or in the decision to submit the article for publication.

## DECLARATION OF INTERESTS

The views expressed in this presentation are the sole responsibility of the presenter and do not necessarily reflect the views, opinions, or policies of the Uniformed Services University of the Health Sciences, the Department of Defense, the Walter Reed National Military Medical Center, the Department of the Navy, Army, Air Force, the United States Government, or the Henry M. Jackson Foundation for the Advancement of Military Medicine, Inc. (HJF). Several of the authors are U.S. Government employees. This work was prepared as part of their official duties. Title 17 US.C. § 105 provides that “Copyright protection under this title is not available for any work of the United States Government”. Title 17 US.C. § 101 defines a U.S. Government work as a work prepared by a military service member or employee of the U.S. Government as part of that person’s official duties. Mention of trade names, commercial products, or organizations does not imply endorsement by the U.S. Government. The study protocol was approved by the USUHS Institutional Review Board in compliance with all applicable federal regulations governing the protection of human subjects.

## CONFLICT OF INTEREST

S.P. and T.H.B. report that the USU IDCRP, a U.S. Department of Defense DoD Institution, and the HJF were funded under a Cooperative Research and Development Agreement to conduct an unrelated phase III COVID-19 monoclonal antibody immunoprophylaxis trial sponsored by AstraZeneca. The HJF, in support of the USU IDCRP, was funded by the DoD Joint Program Executive Office for Chemical, Biological, Radiological, and Nuclear Defense to augment the conduct of an unrelated phase III vaccine trial sponsored by AstraZeneca. Both trials were part of the USG COVID-19 response. Neither is related to the work presented here.

The remaining authors declare that the research was conducted in the absence of any commercial or financial relationships that could be construed as a potential conflict of interest.

## Notes

### Author Declarations

Ethics committee/IRB of Uniformed Services University gave ethical approval for this work.

