## Supplemental Figures and Tables for "Subclinical SARS-CoV-2 Infections and Endemic Human Coronavirus Immunity Shape SARS-CoV-2 Saliva Antibody Responses"

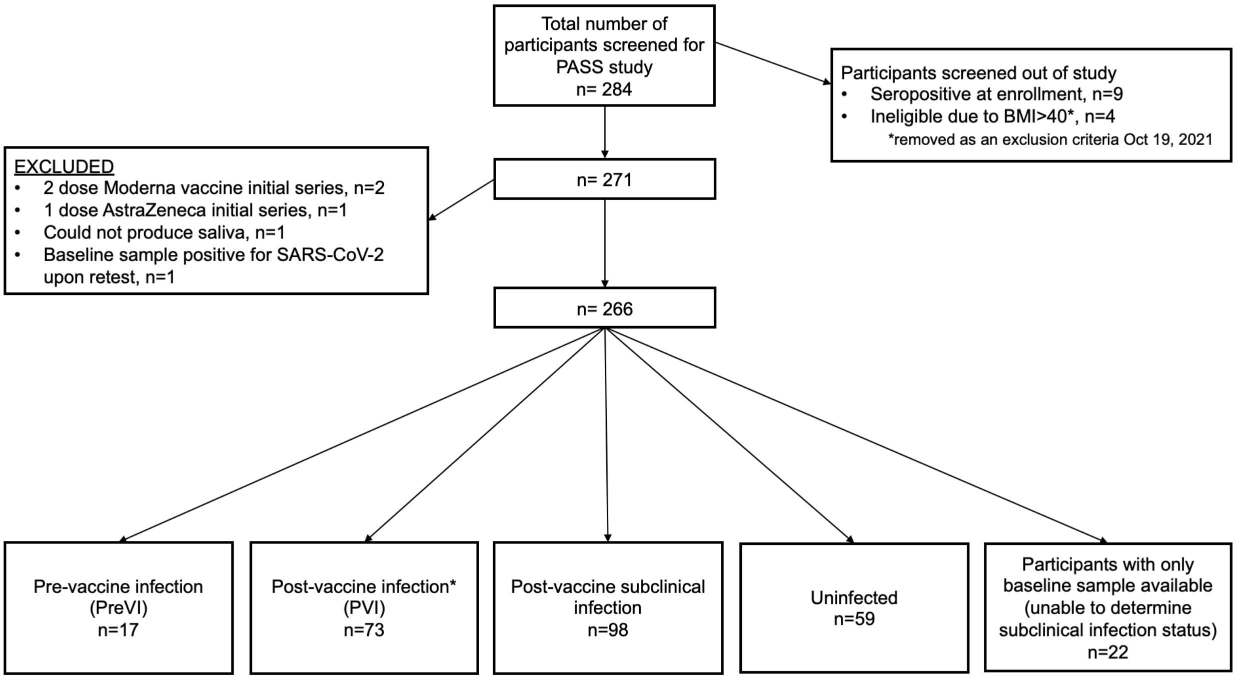


* Includes 3 participants that had pre-vaccine infections and post-vaccine infections

**Supplemental Figure 1. Strobe chart**

**Supplemental Table 1: Study demographics**

| | | |
|---|---|---|
| **Sex** | Female | 185/266 (69.5%) |
| | Male | 81/266 (30.5%) |
| **Age Groups** | 20-29 years old | 37/266 (13.9%) |
| | 30-39 years old | 80/266 (30.1%) |
| | 40-49 years old | 69/266 (25.9%) |
| | 50-59 years old | 55/266 (20.7%) |
| | 60-69 years old | 24/266 (9.0%) |
| | 70-79 years old | 1/266 (0.40%) |
| **Age, median (IQR)** | 41.0 years (33.0, 51.3) | |
| **Race** | Asian | 27/266 (10.2%) |
| | Black | 33/266 (12.4%) |
| | More than one race | 10/266 (3.8%) |
| | Not reported | 6/266 (2.3%) |
| | White | 190/266 (71.4%) |
| **Ethnicity** | Hispanic | 16/266 (6.0%) |
| | Non-Hispanic | 245/266 (92.1%) |
| | Not reported | 5/266 (1.9%) |
| **Occupation** | Nurse | 87/266 (32.7%) |
| | Physician | 69/266 (25.9%) |
| | Occupational/Physical/Speech Therapy | 31/266 (11.7%) |
| | Other clinical support | 28/266 (10.5%) |
| | Laboratory | 21/266 (7.9%) |
| | Behavioral Health | 15/266 (5.6%) |
| | Administrative | 11/266 (4.1%) |
| | Not Reported | 4/266 (1.5%) |

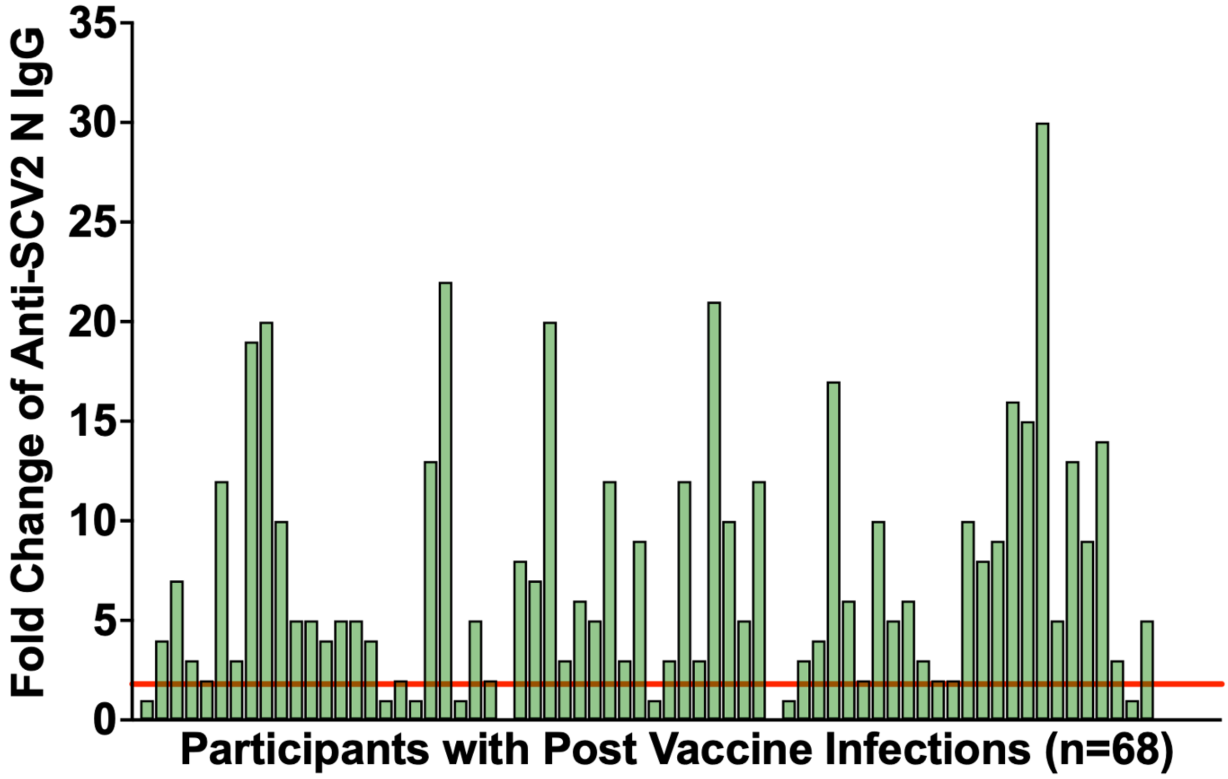


**Supplemental Figure 2. Establishing the saliva anti-SCV2 N IgG threshold for subclinical infections**

Saliva anti-SCV2 N IgG responses for participants with confirmed post-vaccine infections (PVI, n=68). Of this population, 86.8% (n=59) experienced a doubling of anti-SCV2 N IgG compared to baseline levels. This threshold, indicated by the solid red line, was used to determine N positivity indicating a subclinical infection.

**Supplemental Table 2: Categorization of participants by infection and vaccination status**

| | Number of participants (%) N = 266 |
|---|---|
| **Total number of participants with saliva samples collected** | 266 (100.0%) |
| **Participants with only baseline saliva samples collected** | 22 (8.3%) |
| **Participants with at least one positive SARS-CoV-2 test** | 87 (32.7%) |
| Pre-vaccine infection (PreVI) | 17 (6.4% |
| Post-vaccine infection (PVI)* | 73 (26.3%) |
| **Participants with no positive SARS-CoV-2 test during the study** | 157 (59.0%) |
| Subclinical infection | 98 (36.8%) |
| No evidence of any prior infection | 59 (22.2%) |

*PVI group includes 3 participants that were also infected before vaccine

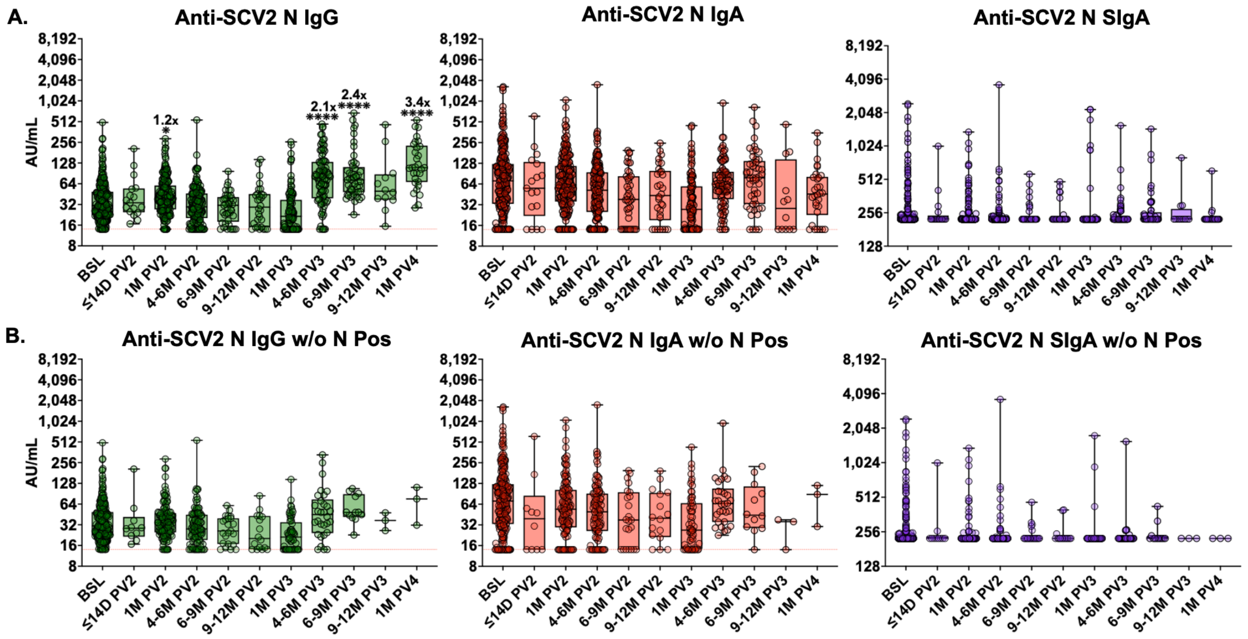


**Supplementary Figure 3: Saliva anti-SCV2 N antibody levels at specific timepoints after vaccination**

**A)** Anti-SCV2 N IgG, IgA and SIgA saliva antibody responses at each time point after vaccination compared to baseline levels in uninfected participants (no positive SARS-CoV-2 test) (BSL n=261, ≤14D PV2 n=17, 1M PV2 n=205, 4-6M PV2 n=175, 6-9M PV2 n=41, 9-12M PV2 n=30, 1M PV3 n=110, 4-6M PV3 n=93, 6-9M PV3 n=49, 9-12M PV3 n=12, 1M PV4 n=30).

**B)** Anti-SCV2 N IgG, IgA and SIgA saliva antibody responses at each time point after vaccination compared to baseline levels after removal of participants with a doubling of anti-SCV2 N IgG in saliva (subclinical infection) as well as those with prior documented SARS-COV-2 infection (BSL n=261, ≤14D PV2 n=10, 1M PV2 n=129, 4-6M PV2 n=105, 6-9M PV2 n=23, 9-12M PV2 n=16, 1M PV3 n=61, 4-6M PV3 n=30, 6-9M PV3 n=12, 9-12M PV3 n=3, 1M PV4 n=3). Comparisons made using log-transformed data and Kruskal-Wallis analysis with Dunn's multiple comparison test. Fold-change in geometric mean compared to baseline is indicated in the text above box and whisker plots. * $p < 0.05$; ** $p < 0.01$; *** $p < 0.001$; **** $p < 0.0001$. (BSL = baseline, D = days, M = months, PV2 = post-2$^{nd}$ vaccine dose, PV3 = post-3$^{rd}$ vaccine dose, PV4 = post-4$^{th}$ vaccine dose). Red dotted line represents lower limit of the assay.

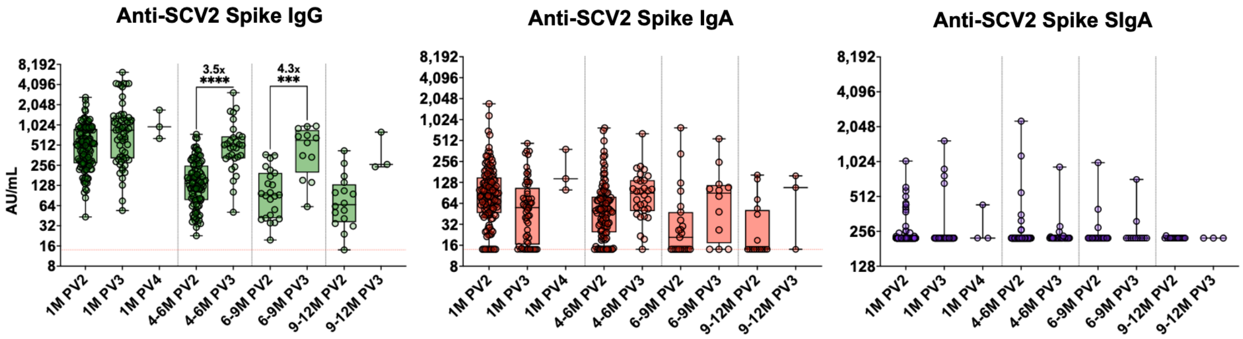


**Supplemental Figure 4: Boosting increases magnitude and durability of saliva anti-SCV2 spike IgG, but does not result in increases to saliva IgA or SIgA**

Saliva antibody responses are compared at 1 month after the 2$^{nd}$ doses of vaccine (1M PV2, n=129), 3rd dose (1M PV3, n=61) and 4th dose (1M PV4, n=3), 4-6 months after 2$^{nd}$ and 3$^{rd}$ doses (4-6M PV2, n=104 and 4-6M PV3, n=30), 6-9 months after 2$^{nd}$ and 3$^{rd}$ doses (6-9M PV2, n=23 and 6-9M PV3,  n=12), and 9-12 months after 2$^{nd}$ and 3$^{rd}$ doses (9-12M PV2, n=16 and 9-12M PV3, n=3). All samples tested were from participants with no prior documented or subclinical SARS-CoV-2 infection at the timepoint tested. Comparisons made using log-transformed data and Kruskal-Wallis analysis with Dunn's multiple comparison test. Fold-change in geometric mean compared to baseline levels is indicated in the text above box and whisker plots. * $p < 0.05$; ** $p < 0.01$; *** $p < 0.001$; **** $p < 0.0001$. Red dotted line represents lower limit of the assay.

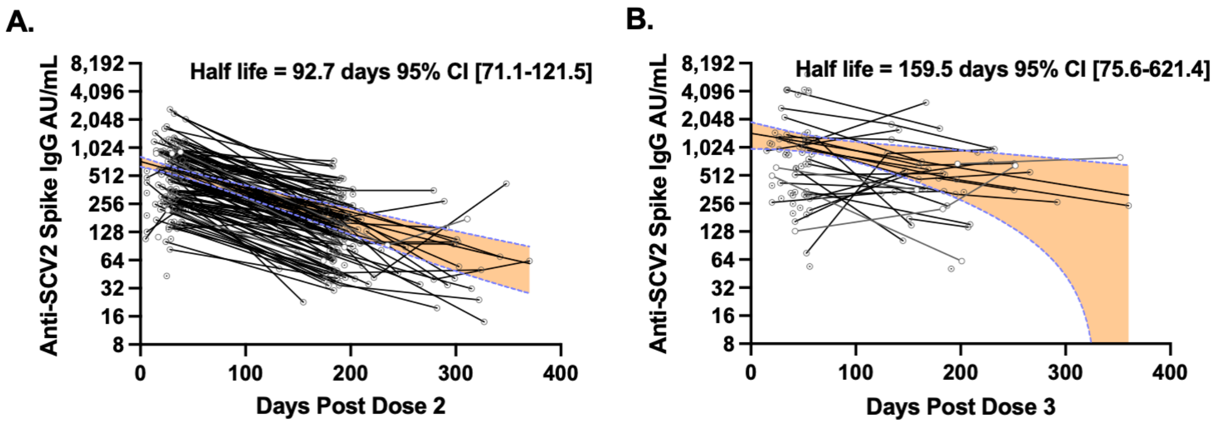


**Supplemental Figure 5: Half-life of saliva anti-SCV2 spike IgG is increased after three doses compared to two doses**

Half-life of saliva anti-SCV2 spike IgG after the 2nd **A)** or the 3rd **B)** dose of mRNA vaccine calculated by one-phase decay modeling. All samples tested were from participants with no prior documented or subclinical SARS-CoV-2 infection at the timepoint tested.

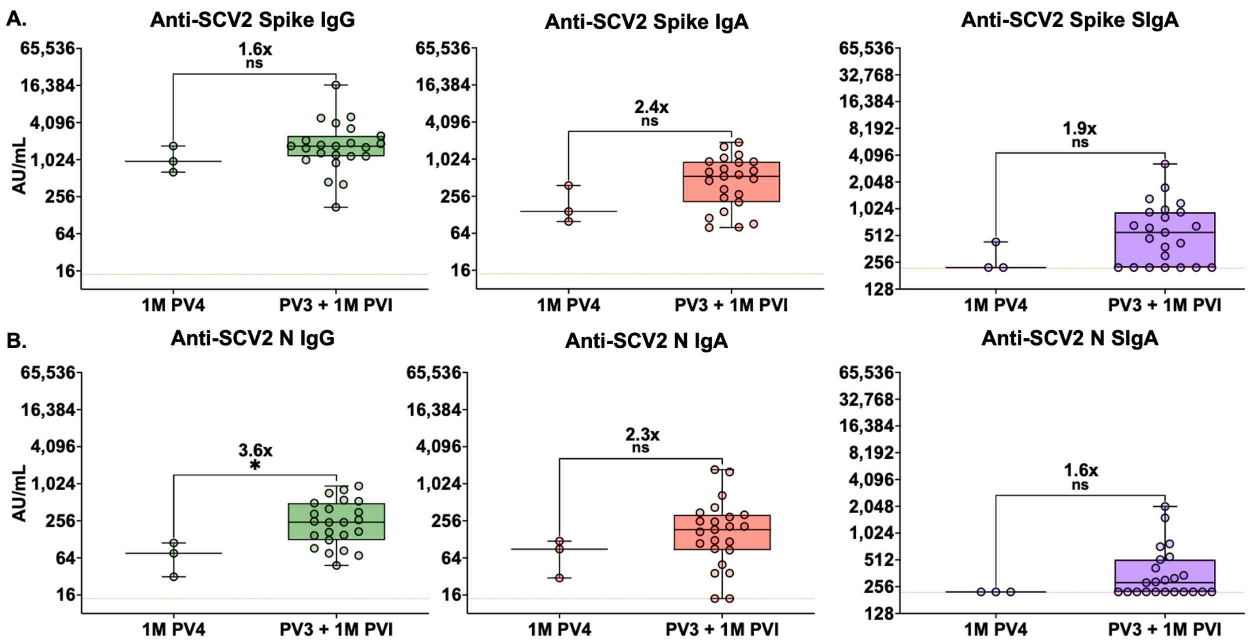


**Supplemental Figure 6: Hybrid immunity induces higher anti-SCV2 spike IgA and SIgA levels than 4 vaccine doses**

**A)** Anti-SCV2 spike and **B)** anti-SCV2 N IgG, IgA and SIgA responses in saliva 1 month after the 4th vaccine dose in uninfected participants (1M PV4, n=3) compared with responses 1 month a post-vaccine infection that occurred after 3 initial vaccine doses (PV3 + 1M PVI, n=23). Analysis performed on log-transformed data using Mann-Whitney test. Fold-change in geometric mean is indicated in the text above box and whisker plots. ns, not significant; * $p < 0.05$; ** $p < 0.01$; *** $p < 0.001$; **** $p < 0.0001$. Red dotted line represents lower limit of the assay.

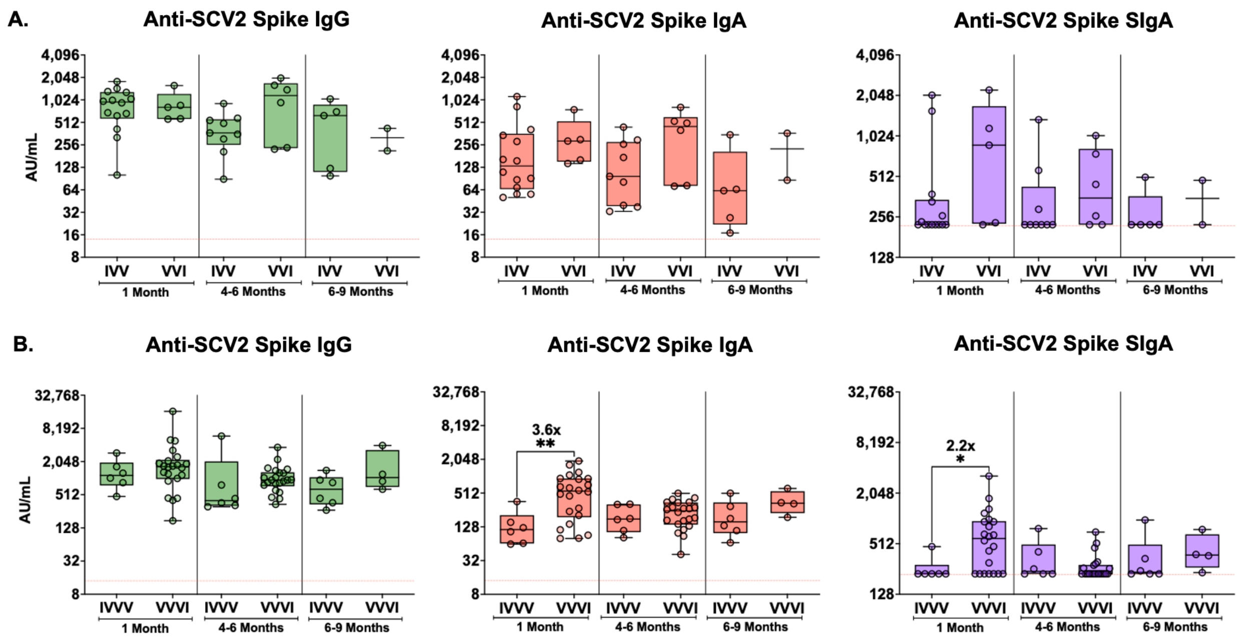


**Supplemental Figure 7: Infection after vaccination results in higher levels of saliva anti-SCV2 spike IgA than vaccination after infection**

**A)** Saliva antibody levels in participants that were infected prior to 2nd vaccine dose (IVV) obtained at 1 month (n=14), 4-6 months (n=9) and 6-9 months (n=5) after the 2$^{nd}$ vaccine dose compared to saliva antibody levels measured in individuals at 1 month (n=5), 4-6 months (n=6) and 6-9 months (n=2) after SARS-CoV-2 infection that occurred following 2 vaccine doses (VVI). **B)** Saliva antibody levels from participants that were infected prior to three vaccine doses (IVVV) obtained at 1 month (n=6), 4-6 months (n=6), and 6-9 months (n=6) after three vaccine doses compared to saliva antibody levels measured in individuals at 1 month (n=22), 4-6 months (n=22) and 6-9 months (n=4) after SARS-CoV-2 infection that occurred following 3 vaccine doses (VVVI). Saliva antibody levels were compared with Kruskal-Wallis and Dunn's multiple comparison test using log-transformed data. Fold-change in geometric mean is indicated in the text above box and whisker plots. * $p < 0.05$; ** $p < 0.01$; *** $p < 0.001$; **** $p < 0.0001$. Red dotted line represents lower limit of the assay.

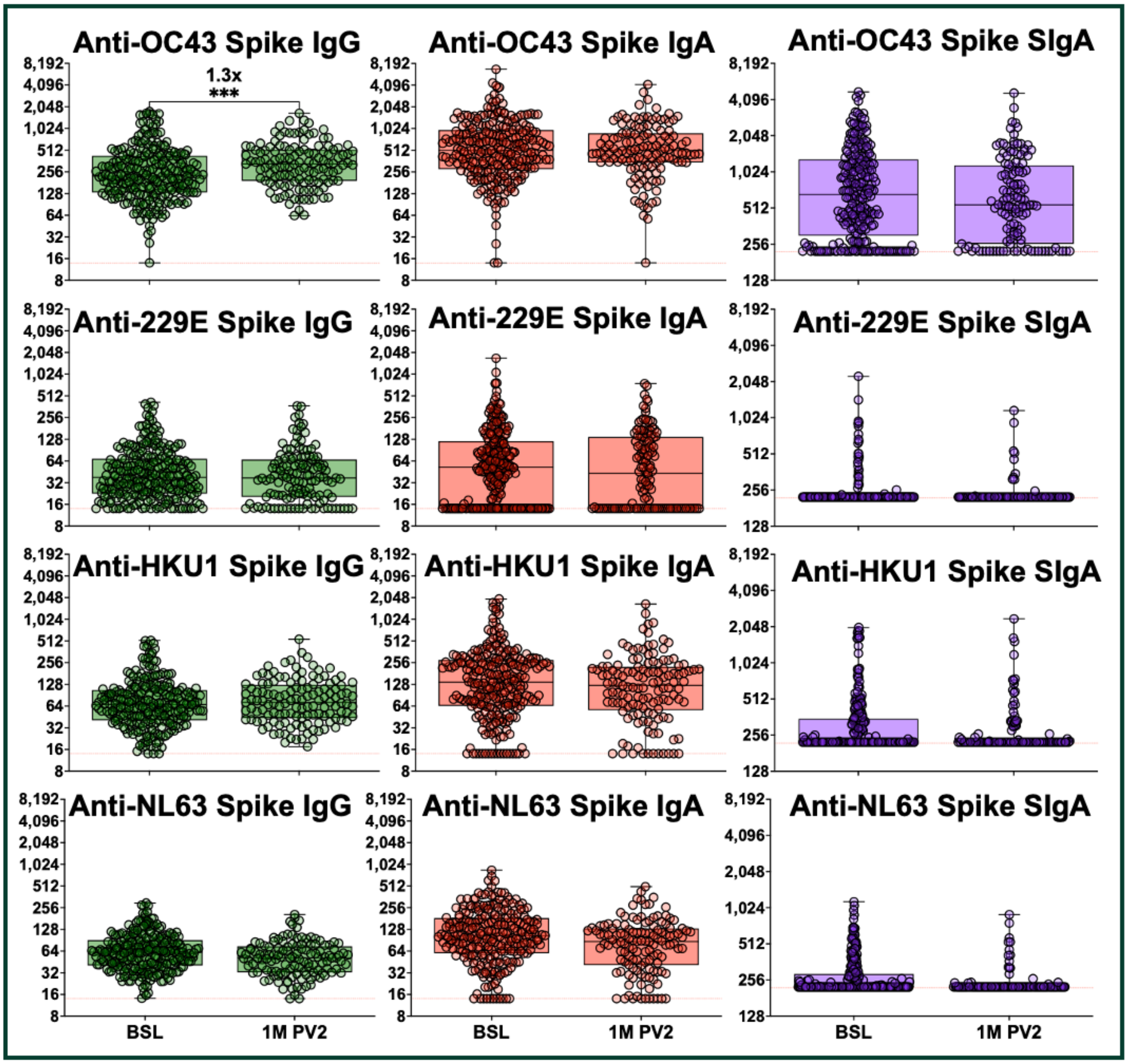


**Supplemental Figure 8: Vaccination results in boosting of IgG against the spike protein of OC43**

Saliva IgG, IgA and SIgA levels against the spike protein of the alpha coronaviruses, NL63 and 229E, and the beta coronaviruses HKU1 and OC43 at baseline (BSL, n=261) and 1 month after the 2nd COVID-19 mRNA vaccine dose (1M PV2, n=129). Comparisons were made using log-transformed data and analyzed with a Mann-Whitney test. Fold-change in geometric mean compared to baseline levels are indicated in the text above box and whisker plots. * $p < 0.05$; ** $p < 0.01$; *** $p < 0.001$; **** $p < 0.0001$. Red dotted line represents lower limit of the assay.

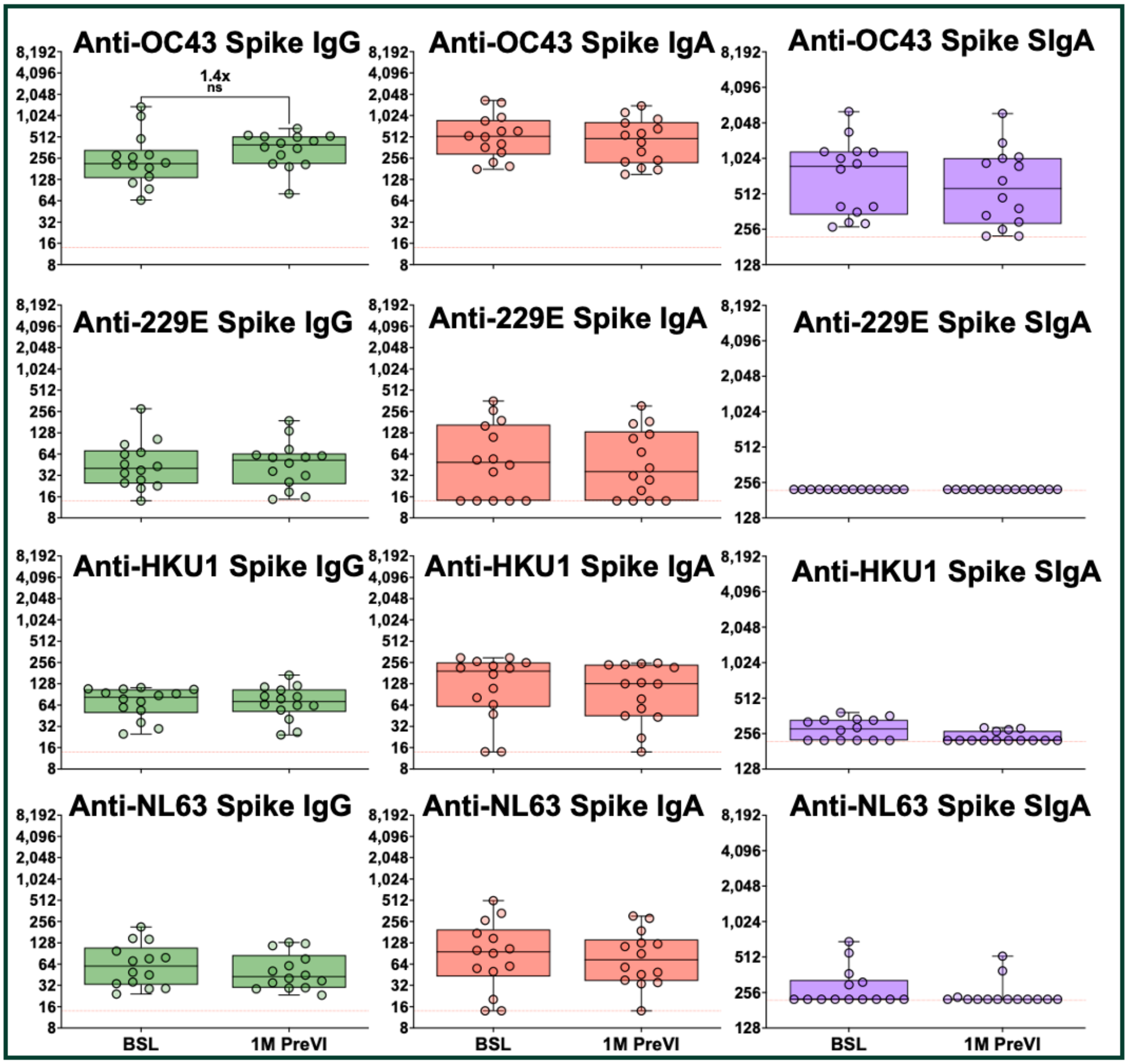


**Supplemental Figure 9: Infection also results in boosting of IgG against the spike protein of OC43**

Saliva IgG, IgA and SIgA levels against the spike protein of the alpha coronaviruses, NL63 and 229E, and the beta coronaviruses HKU1 and OC43 at baseline (BSL) and 1 month after infection (1M PreVI, n=14). These responses were compared to the paired saliva IgG, IgA and SIgA antibody levels against the spike protein of each HCoV at 1 month after infection in these participants who were infected prior to vaccination. Comparisons were made using log-transformed data and analyzed with a Wilcoxson paired test. Fold-change in geometric mean compared to baseline levels are indicated in the text above box and whisker plots. ns, not significant; * $p < 0.05$; ** $p < 0.01$; *** $p < 0.001$; **** $p < 0.0001$. Red dotted line represents lower limit of the assay.
